# Genomic Sequencing of SARS-CoV-2 in Rwanda: evolution and regional dynamics

**DOI:** 10.1101/2021.04.02.21254839

**Authors:** Yvan Butera, Enatha Mukantwari, Maria Artesi, Jeanne D’Arc Umuringa, Áine Niamh O’Toole, Verity Hill, Stefan Rooke, Samuel Leandro Hong, Simon Dellicour, Onesphore Majyambere, Sebastien Bontems, Bouchra Boujemla, Josh Quick, Paola Cristina Resende, Nick Loman, Esperance Umumararungu, Alice Kabanda, Marylin Milumbu Murindahabi, Patrick Tuyisenge, Misbah Gashegu, Jean Paul Rwabihama, Reuben Sindayiheba, Djordje Gikic, Jacob Souopgui, Wilfred Ndifon, Robert Rutayisire, Swaibu Gatare, Tharcisse Mpunga, Daniel Ngamije, Vincent Bours, Andrew Rambaut, Sabin Nsanzimana, Guy Baele, Keith Durkin, Leon Mutesa, Nadine Rujeni

## Abstract

The severe acute respiratory syndrome coronavirus 2 (SARS-CoV-2), responsible for coronavirus disease 19 (COVID-19), is a single-stranded positive-sense ribonucleic acid (RNA) virus that typically undergoes one to two single nucleotide mutations per month. COVID-19 continues to spread globally, with case fatality and test positivity rates often linked to locally circulating strains of SARS-CoV-2. Furthermore, mutations in this virus, in particular those occurring in the spike protein (involved in the virus binding to the host epithelial cells) have potential implications in current vaccination efforts. In Rwanda, more than twenty thousand cases have been confirmed as of March 14^th^ 2021, with a case fatality rate of 1.4% and test positivity rate of 2.3% while the recovery rate is at 91.9%. Rwanda started its genomic surveillance efforts, taking advantage of pre-existing research projects and partnerships, to ensure early detection of SARS-CoV-2 variants and to potentially contain the spread of variants of concern (VOC). As a result of this initiative, we here present 203 SARS-CoV-2 whole genome sequences analyzed from strains circulating in the country from May 2020 to February 2021. In particular, we report a shift in variant distribution towards the newly emerging sub-lineage A.23.1 that is currently dominating. Furthermore, we report the detection of the first Rwandan cases of the VOCs, B.1.1.7 and B.1.351, among incoming travelers tested at Kigali International Airport. We also discuss the potential impact of COVID-19 control measures established in the country to control the spread of the virus. To assess the importance of viral introductions from neighboring countries and local transmission, we exploit available individual travel history metadata to inform spatio-temporal phylogeographic inference, enabling us to take into account infections from unsampled locations during the time frame of interest. We uncover an important role of neighboring countries in seeding introductions into Rwanda, including those from which no genomic sequences are currently available or that no longer report positive cases. Our results point to the importance of systematically screening all incoming travelers, regardless of the origin of their travels, as well as regional collaborations for durable response to COVID-19.

## Introduction

The coronavirus disease 2019 (COVID-19) due to severe acute respiratory syndrome coronavirus 2 (SARS-CoV-2) continues to impose a heavy death toll globally and represents a major global health challenge. Real-time whole-genome sequencing provides invaluable insights on the pandemic’s transmission dynamics and enables effective surveillance. Moreover, genomic data provides useful information required for the ongoing development of vaccines, therapeutics and diagnostic tools. Analysis of SARS-CoV-2 mutations is particularly crucial when these affect epitopes involved in the induction of host immune responses as they may lead to immune evasion, with potential implications for vaccine (and immunotherapy) efficacy.

The global SARS-CoV-2 lineage nomenclature has already been proposed with A and B as the initial phylogenetic lineages ^1^, followed by a number of sub-lineages. Emerging SARS-CoV-2 variants are circulating globally and a number of variants of concern (VOC) have been reported such as the B.1.1.7 VOC (also known as 20I/501Y.V1 or VOC 202012/01), which is characterized by twenty-three mutations (thirteen non-synonymous mutations, four deletions and six synonymous mutations), is associated with higher transmissibility ^2^ and increased mortality ^3,4^; and the B.1.351 VOC (known as 20H/501Y.V2), which emerged independently of B.1.1.7, shares some mutations with the B.1.1.7 VOC and has recently also been associated with low vaccine efficacy in South Africa^5^. Another VOC known as P.1 was first identified in Brazil and is characterized by seventeen unique mutations, including three in the receptor binding domain of the spike protein ^6^.

In Rwanda, the first case of SARS-CoV-2 was confirmed in the capital city, Kigali, on March 14^th^, 2020 following a series of testing at the borders and the Kigali International Airport, (KIA), and was linked to incoming travelers from Mumbai, India. Subsequently, a countrywide total lockdown, coupled with strict prevention measures including contact tracing, was enforced for nearly two months aiming to contain the spread of the virus. From May 2020, lockdown restrictions were lifted progressively, a number of commercial activities resumed and the KIA reopened on the 1^st^ of August 2020. However, despite continued massive testing ^7^, contact tracing, hotspot mapping and preventive measures ^8^, the number of cases continued to increase, mainly associated with cross-border land travels through truck drivers ^9^ and imported cases. This culminated in a ‘first wave’ of local transmission between July and September 2020. Additional containment measures led to the decline of cases until November 2020 when schools and most activities resumed. Since December 2020, another ‘wave’ of infections hit the country, peaking in January-February 2021. As a result, new movement restrictions were enforced, including a total lockdown in the capital city and a seven days’ quarantine for international travelers in addition to two negative polymerase chain reaction (PCR) tests, one pre-departure and another one upon arrival.

In this study, we describe the dynamics of transmission based on genomic analysis of isolates from the first and second waves of the epidemic in Rwanda. In particular, we highlight a shift from ancestral dominant B.1.380 lineage in the early stages of local transmission to a new lineage, A.23.1, that is currently dominating throughout the country. Combining the collected genomic sequence data with individual travel histories to perform travel history-aware phylogeographic inference, we infer introductions into Rwanda from all of its surrounding countries including those from which no genomic sequences are available. Given the importance of these findings on regional surveillance of SARS COV-2, we emphasize the need for strengthening genomic surveillance at the country’s points of entry following the detection of the first cases of the B.1.1.7 and B.1.351 VOCs among travelers arriving at KIA.

## Results

### Patient characteristics

As of the 10th February 2021, a total of 16,865 cases have been confirmed in the country and the sequences analyzed represent 1.2% of the total confirmed cases. The proportion of daily confirmed cases versus the number of sequences taken is illustrated in Figure 1. We sampled a total of 203 cases (reflecting the national screening efforts at points of entry and emerging hotspots) with an average age of 36.7 years, of whom 131 were males and 70 females (and two unknowns) in this study. Of these, location data was available for 152 individuals, of whom 99 lived in Kigali while others were living in different districts of the country (Figure 1). Significant efforts were made to obtain associated metadata for all cases, with specific attention to individual travel history data, as these may shed light on the origins of viral variants introduced from neighboring countries. Of the 203 cases, 28 had recorded travel history (mainly sampled at the airport and other points of entry through the national monitoring and testing efforts) from Tanzania (6), Kenya (4), Demographic Republic of Congo (3), Uganda (3), United States of America (2), United Arab Emirates (2), South Sudan (1), Italy (1), Morocco (1), Senegal (1), Canada (1), China (1), Gabon (1), and Burundi (1). We show the travel cases from neighboring countries in Figures 2 and 3 and we provide the GISAID accession identifiers associated with these travel cases in Supplementary Table S1.

**Figure 1.**
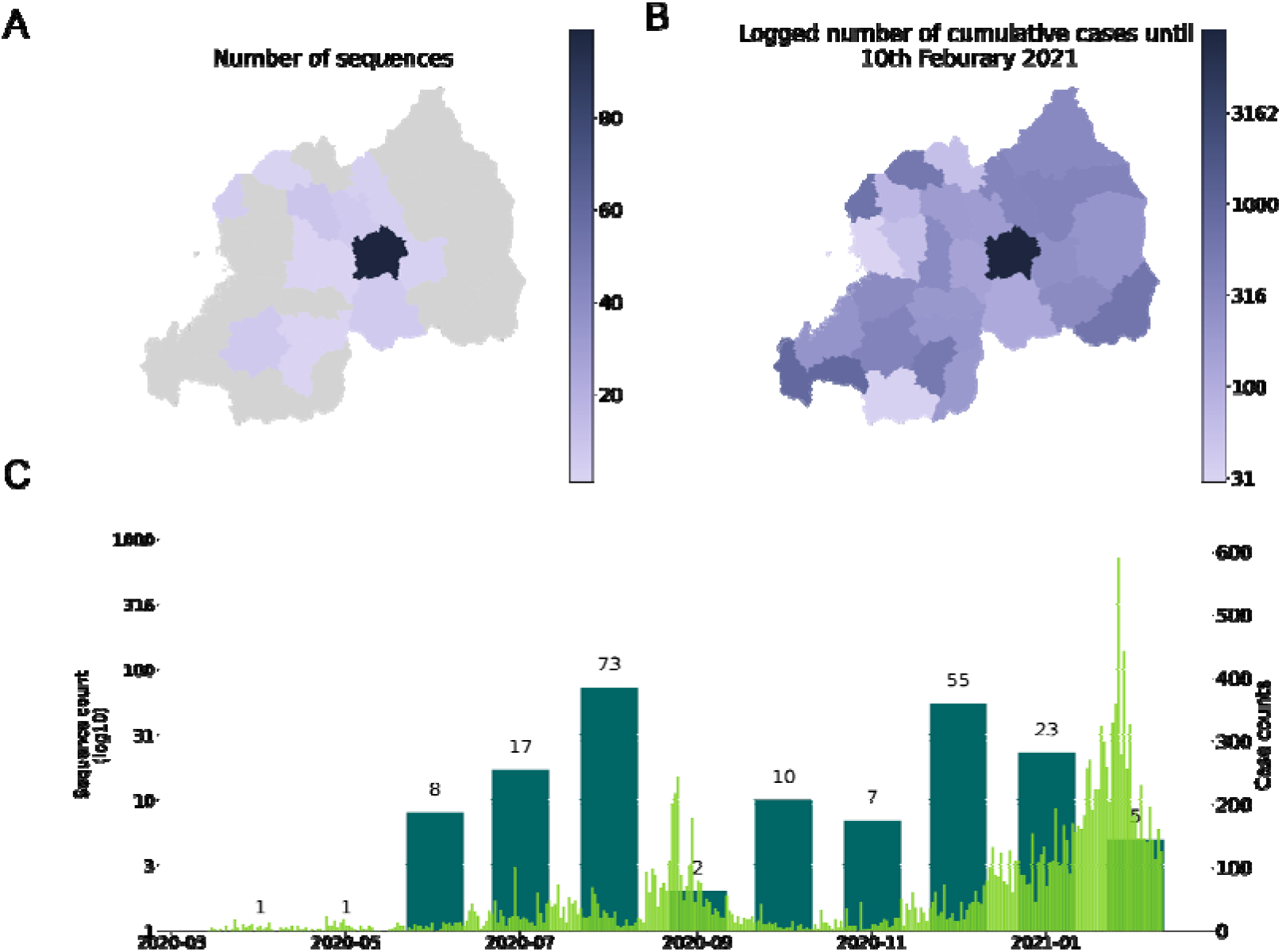
Comparison of the number of sequences taken and case counts over time and space. A) shows the number of sequences in this study by District of residence; B) log-transformed number of cumulative cases by district until the 10^th^ of February, 2021; C) time series of month of sequence collection date (thicker bars), with thinner bars the daily new cases reported nationally until the 10^th^ of February, 2021.

**Figure 2.**
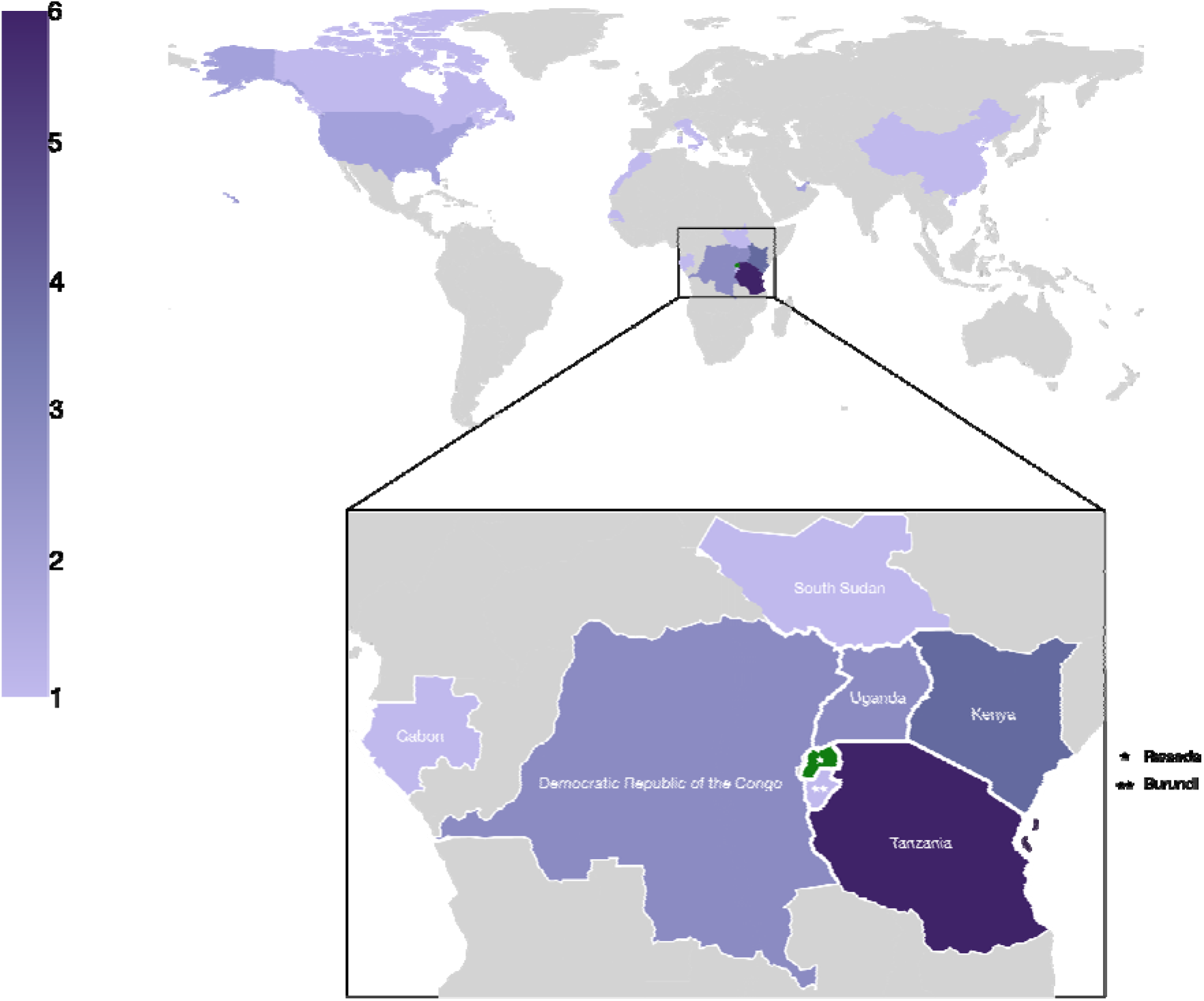
Map showing the number of sequences with recorded travel history per country (n=28). While there is travel into Rwanda recorded from across the world, most cases are from neighboring countries, notably Tanzania (6), Kenya (3), Demographic Republic of Congo (3), Uganda (3), South Sudan (1), Gabon (1), and Burundi (1).

**Figure 3.**
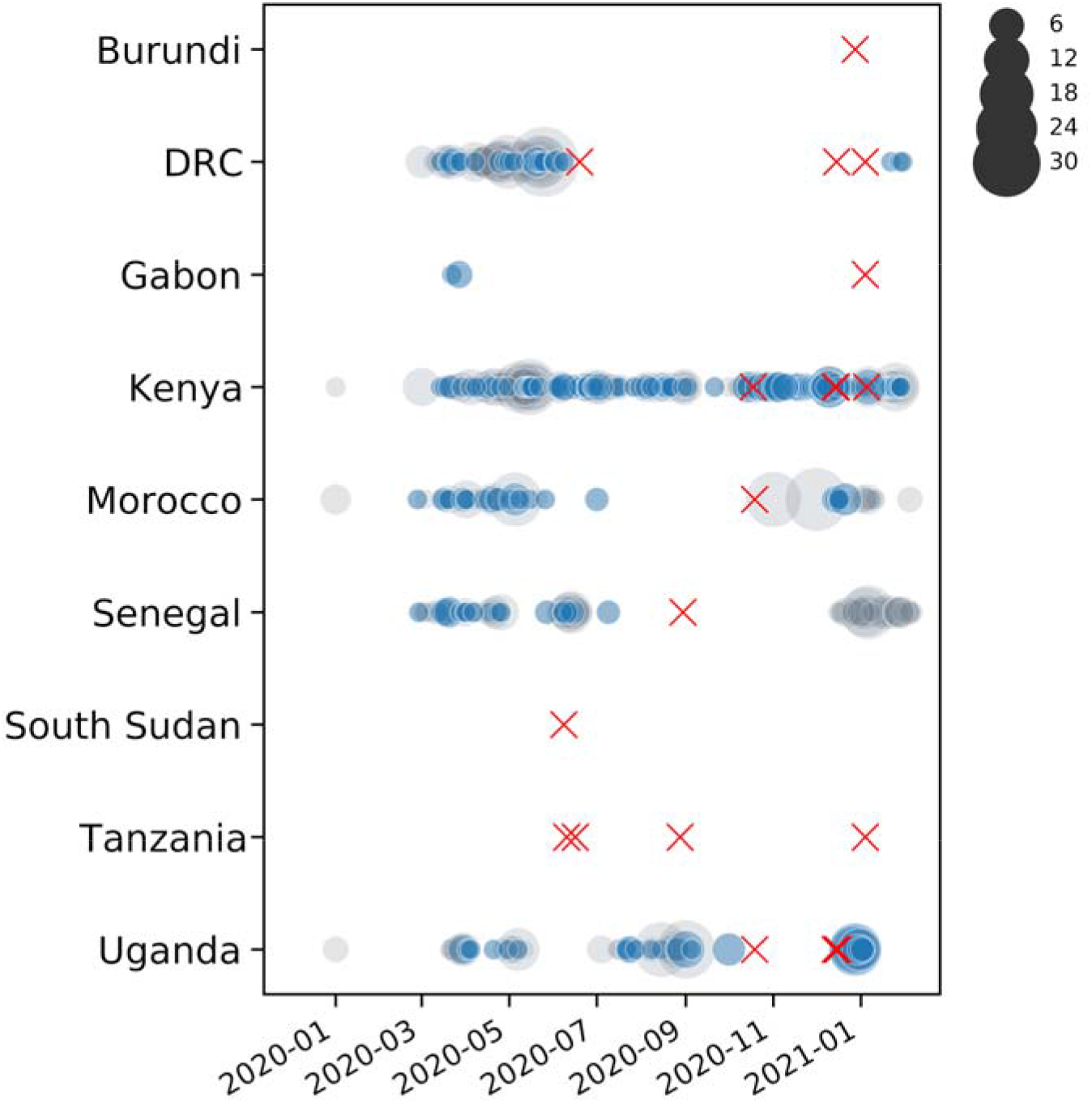
Availability of whole genome sequences for African countries from which travelers entered Rwanda. Grey circles denote the number of sequences available in GISAID for each country on a given date. Blue circles correspond to the number of sequences included in our analyses. Red crosses mark the collection dates of Rwandan sequences with travel history from the respective countries. Although few to no sequences are available from Burundi, Gabon, South Sudan and Tanzania, these travel history data point to SARS-CoV-2 lineages circulating in these countries, to the extent that returning travelers from these countries import those lineages into Rwanda.

### Phylogenetic analysis

The genomes were analyzed using the Pangolin module. Overall, the majority of the SARS-CoV-2 sequences in Rwanda belong to two distinct lineages, A.23.1 and B.1.380. However, the dynamics of their distribution changed over time (Figure 4). Indeed, the early stages of local transmission were characterized by circulation of a dominant B.1.380 lineage, which has only been observed in Rwanda and Uganda. The (limited) diversity of the viral strains observed in the period of May to July 2020 are most likely early imports from Europe and Asia before suppressive measures (such as the countrywide lockdown and the airport closure) were enforced. Nevertheless, an increased strain diversity is observed from the period August-October 2020, most likely reflecting introductions through cross-border land travels for goods and cargo^9^.

**Figure 4.**
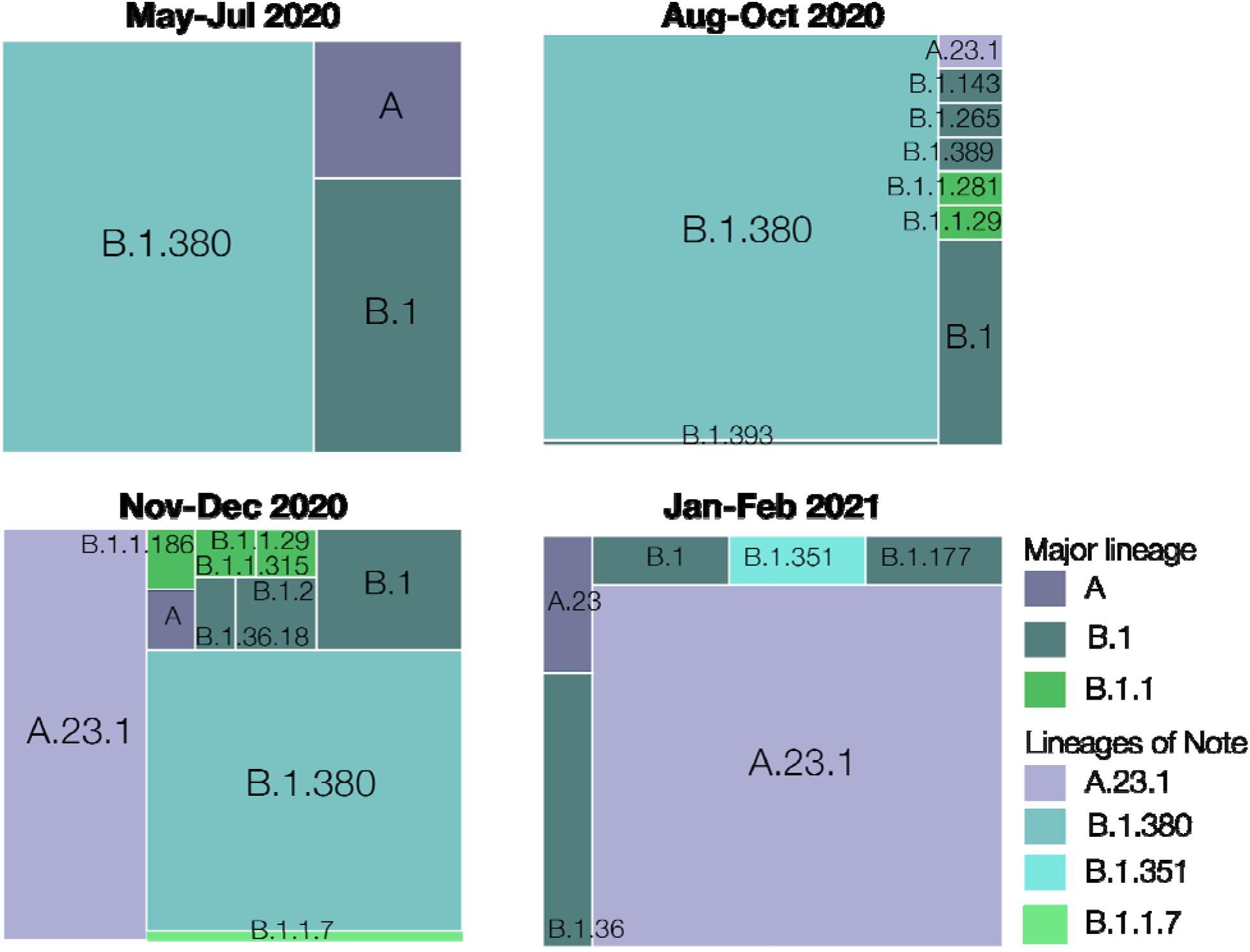
Lineage diversity sampled in Rwanda across four time points: May-Jul 2020 (n=28), Aug-Oct 2020 (n=86), Nov-Dec 2020 (n=74), Jan-Feb 2021 (n=28). Lineage B.1.380, a Rwanda-specific lineage, dominated the sampled diversity during the first wave. Lineage A.23.1 first appeared in Rwanda in October 2020 and quickly became a significant proportion of the sampled SARS-CoV-2 genome sequences. More recently, we detected and sequenced single cases of the B.1.1.7 and B.1.351 VOCs associated with incoming travelers from Burundi and the Democratic Republic of the Congo, respectively.

Towards the end of 2020, we observed a dramatic selective sweep, with a new lineage A.23.1 taking over. This sub-lineage, first observed in Uganda late in 2020 was reported to contain at least 4 amino acid changes in the spike protein and amino acid changes in the proteins nsp3, nsp6, ORF8 and ORF8 ^10^. In particular, Bugembe et al^10^ suggest that the Q613H mutation in spike is may be functionally equivalent to the D614G mutation that arose early in 2020 and is associated with increased viral transmissibility ^12^.

Bugembe et al describe a selective sweep across Uganda of this lineage, which is now the dominant lineage circulating as well. Rwandan genome sequencing shows the presence of A.23.1 as early as 2020-10-21 and a dramatic sweep of this lineage was observed from late November (Figure 4). A.23.1 continues to be the dominant lineage within Rwanda up until February 2021. More recently a number of cases associated with travel have been identified as lineages of concern. The first import cases of B.1.1.7 and B.1.351 variants were sampled on 2020-12-28 and 2021-01-04, respectively. Analysis by Volz et al ^2^ suggests that B.1.1.7 is a more transmissible lineage. Importantly, a recent study suggests that B.1.1.7 is not only more transmissible than preexisting SARS-CoV-2 variants, but it may also cause more severe illness, as indicated by a reported higher hazard of death ^4^. Nevertheless, data inclusive of this paper does not report onward transmission of these VOCs.

### Phylogeographic reconstruction accommodating individual travel histories

We made use of publicly available data and the newly sequenced Rwandan SARS-CoV-2 genomes - all available in GISAID ^15,16^ - to infer a time-scaled phylogenetic tree using maximum-likelihood inference (see Methods). This phylogeny enabled us to identify two subtrees with predominantly Rwandan sequences. Both of these subtrees consist of genetically distinct variants, with the larger cluster belonging to lineage B.1.380 (and hence referred to as subtree B.1) and the smaller one to A.23.1 (referred to as subtree A). The considerable difference in sampling dates and genetic distance between the sequences suggests that the currently circulating SARS-CoV-2 Rwandan lineages are a result of at least two independent introduction events that established local transmission. Subtrees A and B.1 have 172 and 218 sequences, and contain a total of 49 and 134 Rwandan sequences, respectively.

To more accurately understand the pattern of SARS-CoV-2 introduction into Rwanda, we performed a Bayesian discrete phylogeographic analysis on subtrees A and B.1. The 172 genomes in subtree A originated from 33 locations, and included all sequences from lineage A.23.1. The 218 genomes in subtree B originated from 37 locations, and included the B.1.380 lineage. In our analysis of both subtrees, we fit a travel history-aware asymmetric discrete-state diffusion process to model the spatial spread between countries. Our phylogeographic reconstructions included a total of 17 sequences with travel history, 11 for the analysis of subtree A and 6 for subtree B.1 (Table 1). Interestingly, some of these sequences have associated travel histories originating from Tanzania (4 in subtree A and 1 in subtree B.1), a country that has not reported any COVID-19 cases since May 8^th^, 2020 ^17^, and also has no publicly available genomes on GISAID. While Burundi and South Sudan have been consistently reporting case numbers, no genomic sequences are available on GISAID from these countries yet. Our phylogeographic reconstructions are able to include those countries as locations with SARS-CoV-2 infections, by exploiting data on infected incoming travelers from those countries.

Figures 5 and 6 show the estimated location-annotated phylogenies that enable to track the geographic spread of SARS-CoV-2 through time for subtrees A and B.1, with focus on the available Rwandan sequences. In our analysis of subtree A (Figure 5), which contains sequences from lineages A.23 and A.23.1, we inferred a minimum number of 22 (HPD 95%: [16-29]) introduction events into Rwanda, with respectively 13 and 4 of these events originating from Uganda and Kenya (Figure 7; Table S2). We found an expected number of two introduction events from Tanzania into Rwanda, corresponding to and being derived from the two arriving traveler cases, as well as single introduction events from South Sudan and China into Rwanda. Figure 5 also shows frequent mixing between Rwanda, Uganda and Kenya, with the latter two estimated to have seeded introductions into Tanzania, from where no genomic sequences are available to date (see Discussion). However, by employing a travel history-aware inference methodology, we are able to confirm the of lineage A.23.1 among travelers from Tanzania, despite the absence of genomic data. In our analysis of subtree B.1, which includes Rwandan lineage B.1.380, we inferred a minimum number of 9 (HPD 95%: [8-12]) introduction events into Rwanda, with 3 of these events originating from Kenya (Figure 7; Table S2). We also found an expected number of 2 introduction events from both Uganda and Italy.

**Figure 5.**
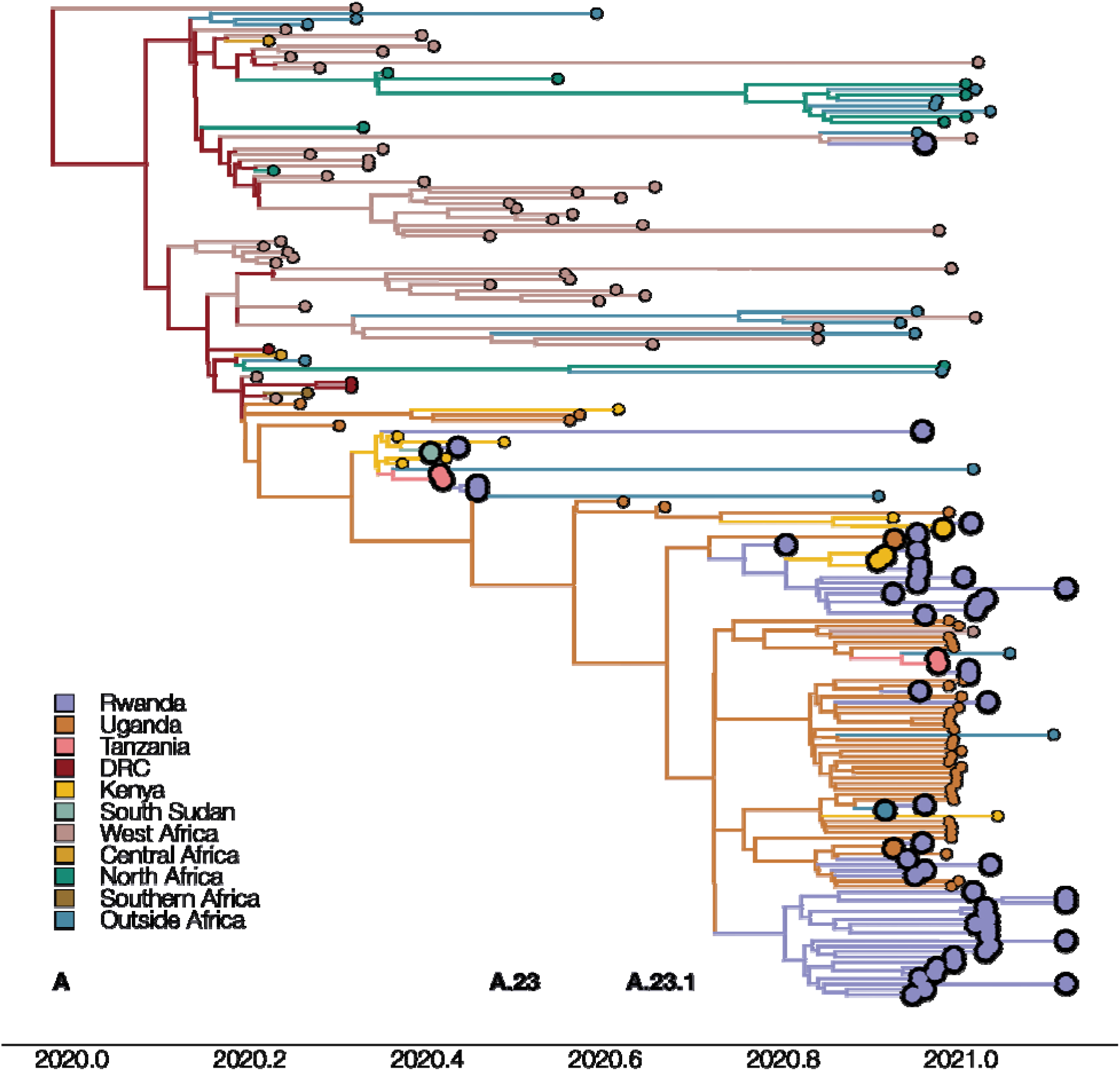
Maximum clade credibility phylogeny for subtree A, representing diversity of lineages A.23 and A.23.1. The phylogeny with associated ancestral locations was inferred using travel history-aware asymmetric discrete state phylogeographic inference. A total of 33 locations were considered in the analysis but are grouped for visualization purposes. The branches in the phylogeny are colored according to the geographical location of the reconstructed ancestral regions. Rwandan sequences are indicated as large tips, colored by associated travel histories (available for 11 of the Rwandan sequences. The travel history-aware phylogeographic reconstruction on subtree A infers frequent mixing between Rwanda, Uganda and Kenya, with the latter seeing introduction events from both Uganda and Rwanda. Both Kenya and Uganda are estimated to have seeded introductions into Tanzania, with the former also seeding an introduction into South Sudan. Importantly, the travel history-aware approach includes (returning infections from) Tanzania in lineage A.23.1, which could not be inferred via other phylogeographic approaches.

**Figure 6.**
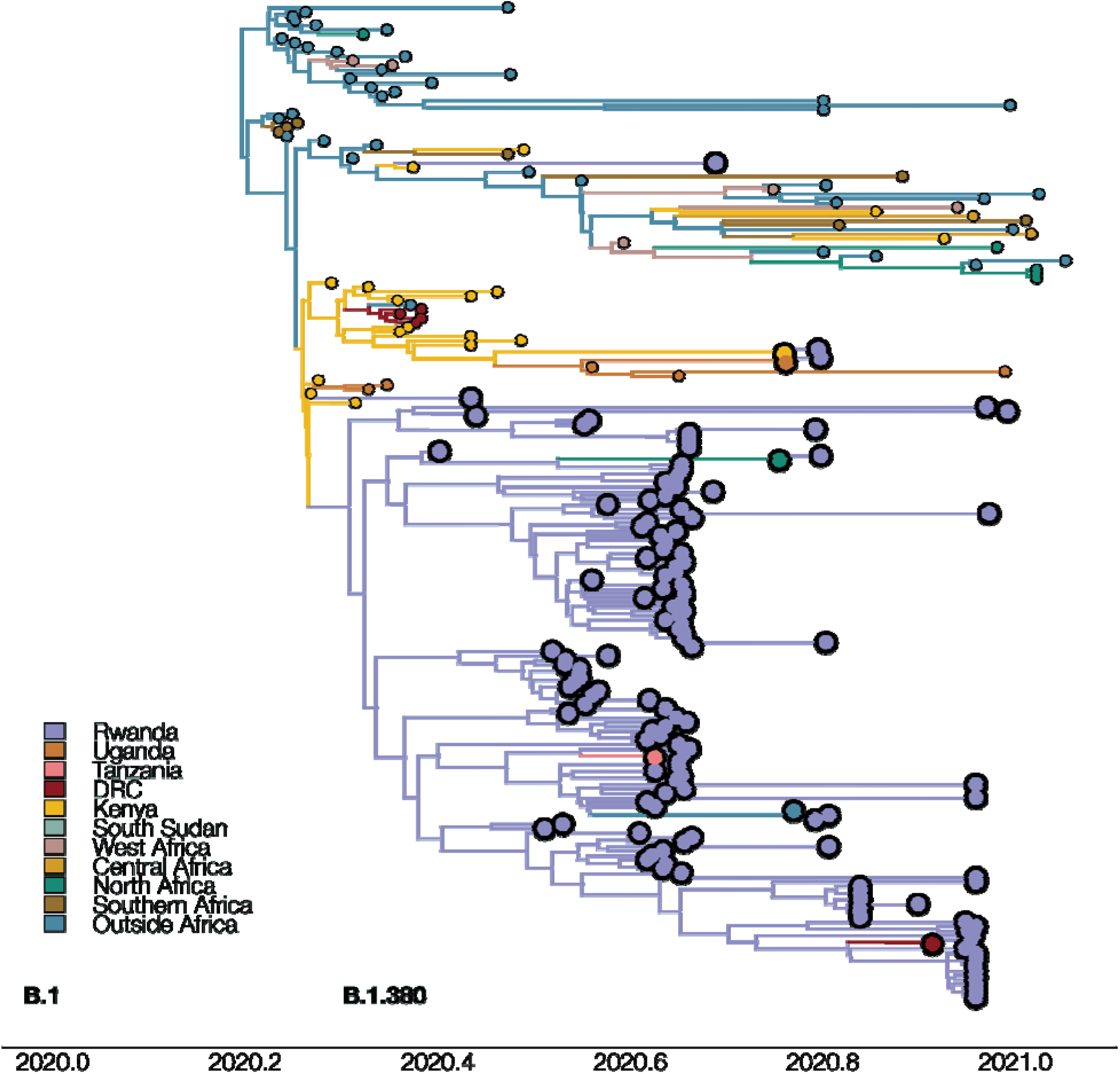
Maximum clade credibility tree for subtree B.1, which includes Rwandan lineage B.1.380. The phylogeny with associated ancestral locations was inferred using travel history aware asymmetric discrete state phylogeographic inference. A total of 37 locations were considered in the analysis. The branches in the phylogeny are colored according to the geographical location of the reconstructed ancestral regions. Rwandan sequences are indicated as large tips, coloured by their associated travel histories. A total of 6 Rwandan sequences with associated travel history are highlighted in this subtree. The travel history-aware phylogeographic reconstruction on subtree B.1 infers a large local transmission cluster in Rwanda (subtree B.1.380). However, by incorporating individual travel histories into the phylogeographic reconstruction, we are able to infer that this subtree does not solely represent local transmission, but also introduction events into Rwanda from Tanzania, Morocco, South Sudan, and the Democratic Republic of the Congo.

**Figure 7.**
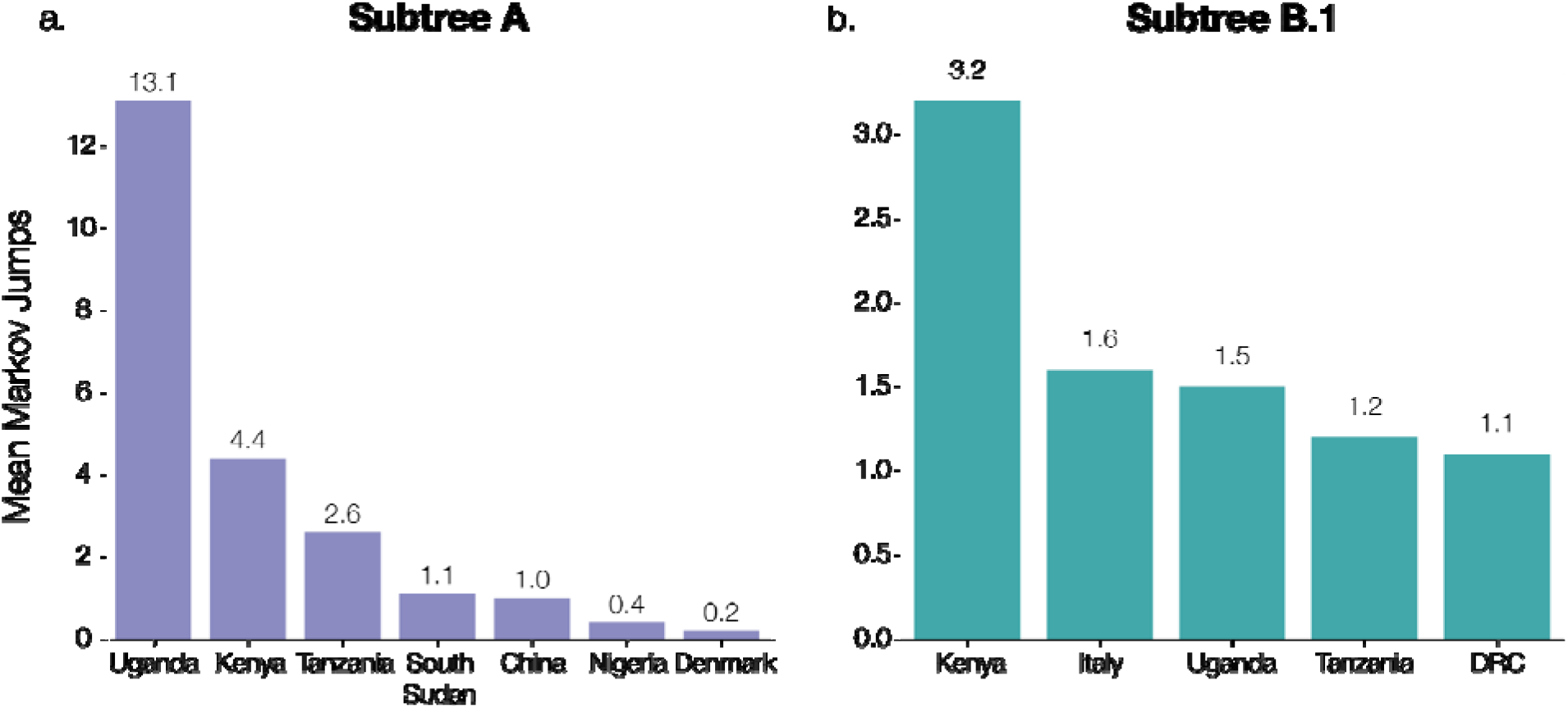
Supported transitions into Rwanda. Mean number of Markov jumps for supported transition rates into Rwanda (Bayes Factor >3) for subtrees A and B.1. Support for these rates was determined using BSSVS with a travel history-aware asymmetric discrete phylogeographic model on both subtrees A and B.1. In both analyses, the majority of introductions into Rwanda was inferred to originate from nearby countries in East Africa, suggesting a substantial exchange of viral lineages between neighboring countries in the region. We refer to Supplementary Table S2 for the Bayes factor support values for these reported transitions.

Using Bayesian stochastic search variable selection (BSSVS), we identified seven statistically supported (Bayes Factor >3) transition routes into Rwanda for subtree A and six for subtree B.1 (Figure 7; Table S2). Our analysis on subtree A showed that Uganda accounted for the majority of SARS-CoV-2 introductions into Rwanda (mean number of Markov jumps: 13.1; 95% HPD: [7-20]), whereas our analysis on subtree B.1 identified Kenya as the main source of SARS-CoV-2 introductions into Rwanda (mean number of Markov jumps: 3.2; 95% HPD: [0-5]).

Consistent with previously published analyses of SARS-CoV-2, we observe that our discrete Bayesian phylogeographic reconstructions resulted in MCC trees of which the internal nodes can be poorly supported, a common phenomenon in SARS-CoV-2 phylogenies (Figures 5 and 6). The considerable uncertainty in phylogenetic clustering results in a variety of diverging phylogeographic histories, which end up not being captured in the MCC trees as these only represent point estimates of the posterior distribution. To this end, we explored the ancestral spatial histories of individual samples of interest using Markov jump trajectory plots ^18,19^ (Figure S3). In the case of subtree A, the travel-aware reconstructions showed four sequences consistently forming two clusters with posterior support >0.9. However, the first two of these four cases correspond to cross-border truck drivers of Tanzanian nationality (sampled on the same day on the same sampling location, i.e. the Rusumo border), with no such metadata available for the other two cases in subtree A. Hence, the two inferred introductions actually correspond to four introduction events from Tanzania into Rwanda, which are clustered together by location in our joint inference, likely as a result of additional samples currently lacking from the border region. Because of this, sequences in each cluster result in nearly identical spatial histories. Figure S3A and S3B show the Markov jump trajectory plots for these two introductions. Overall, we see considerable ambiguity in the ancestral locations prior to Tanzania, as seen by the density of lines landing in “Other” alternate locations. More broadly, we see that in both cases the Rwandan sequences diverged from ancestors in Tanzania, Kenya and Uganda, with considerable uncertainty placed at the root, among the Democratic Republic of the Congo (DRC), Sierra Leone and Mali. The introduction in subtree B, on the other hand, presents us with a different ancestral relation with Tanzania (Figure S3C). Although we also generally observe considerable uncertainty in the ancestral paths, we observe a strong signal for an ancestry in Rwanda prior to the introduction from Tanzania. This would imply a transmission chain starting in Rwanda, spreading into Tanzania, and then being reintroduced into Rwanda. A similar dynamic of outflow and inflow of Rwandan lineages can be seen in the ancestral histories for the sequences with travel history to Morocco, Italy and the DRC (Figure S4). This suggests a bidirectional exchange of SARS-CoV-2 genomes between each of these countries and Rwanda. However, because of the differences in sequencing efforts across the globe, it remains difficult to conclude that such is the case, and the possibility of intermediary locations cannot be discarded. Nonetheless, all spatial trajectory plots imply the presence of SARS-CoV-2 lineages circulating in Tanzania after May 2020. The difference in ancestral histories coupled with the fact that these travel history sequences are genetically distant from each other imply that multiple SARS-CoV-2 lineages have circulated in Tanzania to this day.

In addition, subtree A contains a sequence with travel history to South Sudan. Although over 9,000 COVID cases have been reported up to date ^17^, no genomic sequences are publicly available for South Sudan. The sample tested at arrival in Rwanda presents us with evidence of lineage A having circulated in South Sudan during the months of May and June 2020 (Figure S3D). As expected, the Markov jump trajectory plots for this sample also show considerable uncertainty in the reconstruction of the ancestral locations prior to South Sudan. Regardless, we see some support for ancestry in Kenya, Uganda and the DRC, which provides further evidence for viral transmission between the neighboring countries in the area.

Once introduced in Rwanda, our continuous phylogeographic analysis of SARS-CoV-2 lineages highlight an important inter-connection of those lineages centered around Kigali (Figure S5). Most sequences sampled outside the city appeared to be evolutionarily linked to sequences sampled within this city area, and would then correspond to independent dispersal events from Kigali. However, this phylogeographic pattern, i.e. the central importance of Kigali within the dispersal history of SARS-CoV-2 lineages, might to some extent result from the higher sampling effort within the capital city. Therefore, it is likely that a higher sampling effort outside Kigali would highlight more local transmission.

## Discussion

Here we describe the pattern of transmission of SARS-CoV-2 in Rwanda from May 2020 to February 2021. In particular, we report the spread of a SARS-CoV-2 variant of the A lineage (A.23.1) with notable amino acid changes in the spike protein as well as several non-spike protein changes first detected in Uganda^10^. Indeed, most SARS-CoV-2 sequence diversity in Rwandan strains belong to two distinct lineages: A.23.1 and B.1.380. The latter dominated throughout the early stages of the pandemic before a dramatic shift towards the A.23.1 lineage occurred in November 2020. A similar pattern was observed in neighbouring Uganda as described by Bugembe et al ^10^. The authors describe the new variant as a variant of concern (VOC) in a sense that it shares mutations with the currently known lineage B VOCs such as the changes in key spike protein regions (the furin cleavage site and the 613/614 change). Functional analyses are still needed to determine whether these mutations have effects on transmission rates, immune evasion, vaccine efficacy and/or case fatality rates.

In this study, we reported on the ongoing genomic sequencing efforts in Rwanda, which are complemented with careful collection of associated travel history metadata of incoming travelers. These efforts enabled us to exploit this information by performing joint Bayesian travel history-aware phylogeographic inference on these data. By applying this recently developed approach, we demonstrated considerable contributions of neighbouring countries’ sequence introductions into Rwanda (as well as possible bidirectional exchanges). Of particular interest to this study, we were able to include traveler cases from Tanzania, Burundi and South Sudan while none of these three countries have made any SARS-CoV-2 genomes available throughout the pandemic. According to the data we collected, two infected Rwandan travelers returned from Tanzania on the 16^th^ of June, 2020 and two more on 4^th^ January, 2021. Our findings also complement reports from the WHO (https://www.who.int/news/) that a number of travelers from Tanzania who have travelled to neighboring countries and beyond have tested positive for COVID-19. Incorporating travel history information in phylogeographic analysis can mitigate sampling bias (from unsampled or under-sampled countries) ^18^, although this cannot fully replace the lack of sequences from other countries.

The reported import into Rwanda of 2 VOCs, namely B.1.1.7 and B.1.351, sampled at the Kigali International Airport in late December 2020 and early January 2021 are also particularly interesting. The patient with the B.1.1.7 variant was a Burundian travelling from Burundi while the patient with the B.1.351 variant was a Zimbabwean coming from DRC, suggesting that VOCs may be circulating in neighboring countries. Indeed, although Burundi, South Sudan and Tanzania have currently no SARS-CoV-2 sequences published on GISAID, the DRC has published a total of 371 sequences, of which 3 are VOCs (two B.1.1.7 and one B.1.351), while Kenya has published a total of 686 sequences of which 5 are VOCs (one B.1.1.7 and 4 B.1.351).

Ongoing genomic surveillance in Rwanda indicates additional samples from these VOCs (mostly B.1.351) from travelers sampled at the airport. In an effort to curb the spread of the variants, following the upsurge of cases in November-December 2020, several measures were taken by the Rwandan government including a 7-day quarantine to all incoming passengers followed by a RT-PCR test, in addition to presenting a COVID-19 negative test upon arrival. Furthermore, the capital city of Kigali went through a total lockdown from mid-January to early February 2021, and travels between Districts were prohibited until mid-March 2021. A 7pm to 4am curfew was instituted in early February 2021; public offices were closed and employees were working from their homes. All schools in Kigali were closed as well, and classes were being held online. Cafés and restaurants were only providing take-away services. Churches, public swimming pools and gyms were closed (Office of the Prime Minister - Republic of Rwanda 2021). Such suppression mechanisms (population-wide social distancing, lockdown, school closure, case isolation) have proven the greatest impact (as far as non-pharmaceutical approaches are concerned) in terms of transmission control ^20^. Additionally, all public health facilities received free antigen rapid diagnostic tests for every single person presenting COVID-19 related symptoms. Moreover, a vaccination campaign was initiated in March 2021, with a target to vaccinate all front liners and vulnerable populations (elderly and people with other underlying health conditions) in the first phase. A rapid and efficacious vaccination coverage will ease the social and economic disruptions associated with non-pharmaceutical transmission suppression mechanisms.

These results suggest that neighboring countries play an important role in establishing the circulation of (different strains of) SARS-CoV-2 in Rwanda. However, due to the unevenness in sampling across countries, with several not providing any genomic sequences, additional data would be required to accurately assess the effect of short-distance (e.g. crossing border with neighboring countries) versus long-distance travel in shaping the Rwandan epidemic. Low spread of mutant virus may be partly explained by the fact that Rwanda is relatively less affected by the spread of SARS COV-2 consecutive to several public health measures implemented in light with regular bi-weekly surveillance surveys across the country to monitor the trend of the pandemic.

## Methods

### Study design

This is an in-depth study of SARS-CoV-2 strains that circulate in Rwanda from May 2020 to February 2021, in which we describe the demography and epidemiology of 203 SARS-CoV-2 genomes from collected SARS-CoV-2 positive oropharyngeal swabs. These swabs were obtained from two distinct groups: from individuals residing in different provinces of Rwanda (n=189) and from returning travelers, whose samples were collected at the airport (n=14). All samples were extracted from the biorepository of the National Reference Laboratory (NRL), in Kigali, Rwanda. Samples with a cycle threshold (*Ct*) value below 33 were selected, ensuring a wide geographical representation as well as ports of entry, and case description variables (date and place of RT-PCR test, age, sex, occupation, residence, nationality, travel history) were reported.

### Sequencing

#### RNA Extraction

Ribonucleic acid (RNA) of the virus was extracted from confirmed SARS-CoV-2 positive clinical samples with *Ct* values ranging from 13.4 to 32.7 on a Maxwell 48 device using the Maxwell RSC viral RNA kit (Promega) following a viral inactivation step using proteinase K according to the manufacturer’s instructions.

### SARS-CoV-2 whole genome sequencing

Reverse transcription was performed using SuperScript IV VILO master mix, and 3.3 μl of RNA was combined with 1.2 μl of master mix and 1.5 μl of H2O. This was incubated at 25°C for 10□min, 50°C for 10□min, and 85°C for 5□min. PCRs used the primers and conditions recommended in the nCoV-2019 sequencing protocol (ARTIC Network, 2020) or the 1,200 bp amplicons described by Freed and colleagues ^21^.

Primers from version 3 of the ARTIC Network and the 1,200 bp amplicons were used and were synthesized by Integrated DNA Technologies. Samples were multiplexed using the Oxford Nanopore native barcoding expansion kits 1-12, 13-24, or the native barcoding expansion 96 in combination with the ligation sequencing kit 109 (Oxford Nanopore). Sequencing was carried out on a MinION using R9.4.1 flow cells.

### Genome assembly

The data generated via the Oxford Nanopore Technology (ONT) MinION was processed using the ARTIC bioinformatic protocol (https://artic.network/ncov-2019/ncov2019-bioinformatics-sop.html). Briefly, the FAST5 sequence files were base called and demultiplexed using Guppy 4.2.2 in high accuracy mode, requiring barcodes at both ends of the read. FASTQ reads associated with each sample were filtered and concatenated via the guppy plex module. Consensus SARS-CoV-2 sequences were generated via the ARTIC nanopolish pipeline and assembled for each sample by aligning the respective sample reads to the Wuhan-Hu-1 reference genome (GenBank Accession: MN908947.3) with the removal of sequencing primers, followed by a polishing step using the raw Fast5 signal files. Positions with insufficient genome coverage were masked with N.

### Phylogenetic and phylogeographic analysis

We downloaded all SARS-CoV-2 genomes from the available nextstrain build ^22^ with Africa-focused subsampling (https://nextstrain.org/ncov/africa) on February 23, 2021. These sequences were further complemented to include all 203 Rwandan sequences generated in this study and available on GISAID on February 24, 2021. The 203 Rwandan whole genome SARS-CoV-2 genomes were assigned Pango lineages, as described by Rambaut et al ^1^, using pangolin v2 and pangoLEARN model v2021-02-21 by O’Toole et al (https://github.com/cov-lineages/pangolin). We used Squarify to construct the square treemaps of lineage diversity across three time points (https://github.com/laserson/squarify). We mapped the combined data set against the canonical reference (GISAID ID: EPI_ISL_406801) using minimap2 ^23^ and trimmed the data to positions 265-29,674 and padded with Ns in order to mask out 3’ and 5’ UTRs. We used the resulting alignment to estimate an unrooted maximum-likelihood phylogeny using IQ-TREE v2.1.2 ^24^ using its automated model selection approach that identified the GTR+F+R8 model as best fitting the data. We subsequently calibrated this phylogeny in time using TreeTime ^25^ while estimating the molecular clock and skyline coalescent model parameters and using three SARS-CoV-2 genomes from Wuhan, 2019, as the outgroup.

We went on to perform a discrete Bayesian phylogeographic analysis in BEAST 1.10.5 ^26^ using a recently developed model that is able to incorporate available individual travel history information associated with the newly sequenced Rwandan samples ^18,19^. Exploiting such information can yield more realistic reconstructions of virus spread, particularly when travelers from unsampled or under sampled locations are included to mitigate sampling bias. To this end, and given that it is not feasible to perform such an analysis on the full data set due to the large number of sequences, we selected two subtrees in the overall phylogeny (see Results section) that predominantly consisted of Rwandan sequences, consisting of 172 (subtree A) and 218 sequences (subtree B.1), of which respectively 11 and 6 infected individuals have associated travel history information (see Table S1). We incorporated the collection dates for those sequences into our analyses, and treated the time when the traveler started the return journey to Rwanda as a random variable given that the time of traveling to the sampling location (in Rwanda) was not known (with sufficient precision). We specify normal prior distributions over these 17 random variables informed by an estimate of time of infection and truncated to be positive (back-in-time) relative to sampling date. As in the work of Lemey et al. (2020), we use a mean of 10 days before sampling based on a mean incubation time of 5 days ^27^, and a constant ascertainment period of 5 days between symptom onset and testing ^20^, and a standard deviation of 3 days to incorporate the uncertainty on the incubation time. We retrieved the 172 and 218 sequences from the full alignment and performed joint discrete phylogeographic inference on each resulting data set using BEAST 1.10.5, employing the BEAGLE 3.2.0 high-performance computational library ^28^ to improve performance. For each of these phylogeographic analyses, we make use of Bayesian stochastic search variable selection (BSSVS) to simultaneously determine which migration rates are zero depending on the evidence in the data and efficiently infer the ancestral locations, in addition to providing a Bayes factor test to identify significant non-zero migration rates ^29^. We also estimated the expected number of transitions (known as Markov jumps) ^30,31^ into Rwanda from all other countries in the data set. These analyses ran for a total of 200 and 250 million iterations, respectively, with the Markov chains being sampled every 100,000th iteration, in order to reach an effective sample size (ESS) for all relevant parameters of at least 200, as determined by Tracer 1.7 ^32^. We used TreeAnnotator to construct maximum clade credibility (MCC) trees for each subtree.

To explore the spread of SARS-CoV-2 lineages introduced in Rwanda, we also performed a continuous phylogeographic analysis following a procedure similar to one defined by Dellicour et al. ^33^. Specifically, we used the relaxed random walk (RRW) diffusion model ^34^ available in BEAST 1.10.5 ^26^ to infer the dispersal history of Rwandan lineages along Rwandan clades identified within the two subtree-specific MCC trees that resulted from the discrete Bayesian phylogeographic inference described above. To achieve a sufficient level of spatial precision, the continuous phylogeographic analysis was only based on those sampled genomes for which the Rwandan sector of origin was known, which is the maximal level of spatial precision available for these samples. For each sampled genome associated with this level of sampling precision, which corresponds to 57% of available Rwandan genomes, we retrieved geographic coordinates from a point randomly sampled within its sector of origin. The MCMC chain was run in BEAST 1.10.5 for 30 million iterations and sampled every 10,000^th^ iteration, its convergence/mixing properties were again assessed with Tracer ^32^, and an appropriate number of sampled trees was discarded as burn-in (10%). The resulting sets of plausible trees were used to obtain subtree-specific MCC summary trees using TreeAnnotator, and we then used functions available in the R package “seraphim” ^35^ to extract spatio-temporal information embedded within posterior trees and visualize the continuous phylogeographic reconstructions. Finally, we used the baltic Python library to visualize the phylogenies ^36^.

## Data Availability

All data are available from the corresponding authors upon reasonable request.

## Ethical approval

The study was approved by the Rwanda National Ethics Committee (FWA Assurance No. 00001973 IRB 00001497 of IORG0001100/15April2020) as well as the IRB of the University of Rwanda, College of Medicine and Health Sciences (Approval notice No 325/CMHS IRB/2020).

## Data availability

The reported SARS-CoV-2 genomes are available on GISAID (www.gisaid.org) under the accession numbers EPI_ISL_614763, EPI_ISL_614980, EPI_ISL_615063, EPI_ISL_615064, EPI_ISL_615067, EPI_ISL_615069, EPI_ISL_615071, EPI_ISL_615074, EPI_ISL_615075, EPI_ISL_707711-EPI_ISL_707713, EPI_ISL_ 707771-EPI_ISL_707774, EPI_ISL_707776, EPI_ISL_707777, EPI_ISL_707779, EPI_ISL_707780, EPI_ISL_707783, EPI_ISL_707787-EPI_ISL_707790, EPI_ISL_735436-EPI_ISL_735438, EPI_ISL_735444-EPI_ISL_735448, EPI_ISL_925847-EPI_ISL_925915, EPI_ISL_930567, EPI_ISL_930634, EPI_ISL_930853, EPI_ISL_960227-EPI_ISL_960302, EPI_ISL_1063900-EPI_ISL_1063901, EPI_ISL_1063905, EPI_ISL_1063915, EPI_ISL_1063994, EPI_ISL_1064022, EPI_ISL_1064147-EPI_ISL_1064149, EPI_ISL_1064152-EPI_ISL_1064154, EPI_ISL_1064163-EPI_ISL_1064166, EPI_ISL_1064168,EPI_ISL_1064170, EPI_ISL_1064171

## Contributors

YB, EM: Study design, data collection

MA, YB, BB, EM, JdU: RT-PCR, library preparation, whole genome sequencing

KD, YB, SR: Whole genome sequencing, sequences cleaning and assembling, sequences fast files production

YB, EM, SB, OM, JdU: RNA extraction

PT, RS, MG, RR, EU, AK, MG, OM, RM, SG: Sample selection, data collection

SD, SLH, VH: Spatial and phylogeographic analysis

JR, DG, JS, WN, JQ, MMM, AK, PCR, NL, JPR, SN, TM, DN: Provided technical guidance and review of the paper

GB, AOT: Phylogenetic analysis

VB, JS, WN, GB, AR, SN, KD, MA, LM, NR: Provided scientific and technical guidance, review of the paper

VB, NR, KD, LM: Secured funding and coordinated the whole study

## Acknowledgements

This research was commissioned by the National Institute of Health Research (NIHR) Global Health Research programme (16/136/33) using UK aid from the UK Government (funding to EM and NR through TIBA partnership) and additional funds from the Government of Rwanda through RBC/National Reference Laboratory in collaboration with the Belgian Development Agency (ENABEL) for additional genomic sequencing at GIGA Research Institute-Liege/Belgium. The views expressed in this publication are those of the authors and not necessarily those of the NIHR, the National Institute of Health Research, the Department of Health and Social Care or the Rwandan Government. GB acknowledges support from the Internal Fondsen KU Leuven/Internal Funds KU Leuven (Grant No. C14/18/094) and the Research Foundation—Flanders (“Fonds voor Wetenschappelijk Onderzoek - Vlaanderen,” G0E1420N, G098321N). SLH acknowledges support from the Research Foundation - Flanders (“Fonds voor Wetenschappelijk Onderzoek - Vlaanderen,” G0D5117N). SD is supported by the *Fonds National de la Recherche Scientifique* (FNRS, Belgium). VH was supported by the Biotechnology and Biological Sciences Research Council (BBSRC) [grant number BB/M010996/1]. A.OT is supported by the Wellcome Trust Hosts, Pathogens & Global Health Programme [grant number: grant.203783/Z/16/Z] and Fast Grants [award number: 2236]. AR acknowledges the support of the Wellcome Trust (Collaborators Award 206298/Z/17/Z – ARTIC network) and the European Research Council (grant agreement no. 725422 – ReservoirDOCS).

## Supplementary Materials

**Table S1.** GISAID accession identifiers for the Rwandan sequences in this study for which individual travel history metadata is available. We list the collection date for each GISAID entry, along with the country from which infected travelers returned to Rwanda, and the subtree assignment based on Figure S1.

| Accession ID | Collection date | Travel history | Subtree |
| --- | --- | --- | --- |
| EPI_ISL_707771 | 2020-06-16 | Tanzania | A |
| EPI_ISL_707772 | 2020-06-16 | Tanzania | A |
| EPI_ISL_925865 | 2020-12-17 | China | A |
| EPI_ISL_1063905 | 2020-12-14 | Uganda | A |
| EPI_ISL_925848 | 2020-12-14 | Kenya | A |
| EPI_ISL_925851 | 2020-12-15 | Kenya | A |
| EPI_ISL_1064164 | 2021-01-4 | Tanzania | A |
| EPI_ISL_1064163 | 2021-01-4 | Tanzania | A |
| EPI_ISL_925850 | 2020-12-16 | Uganda | A |
| EPI_ISL_1064154 | 2021-01-5 | Kenya | A |
| EPI_ISL_707712 | 2020-06-8 | South Sudan | A |
| EPI_ISL_735448 | 2020-10-19 | Morocco | B.1 |
| EPI_ISL_960250 | 2020-08-28 | Tanzania | B.1 |
| EPI_ISL_735445 | 2020-10-22 | Italy | B.1 |
| EPI_ISL_925896 | 2020-12-15 | Democratic Republic of the Congo | B.1 |
| EPI_ISL_1063900 | 2020-10-18 | Kenya | B.1 |
| EPI_ISL_707789 | 2020-10-19 | Uganda | B.1 |

**Table S2.**

| From | Subtree | Bayes factor | Posterior probability | Mean Markov jumps |
| --- | --- | --- | --- | --- |
| Uganda | A | 63104 | 1 | 13.1 |
| Tanzania | A | 63104 | >0.99 | 2.6 |
| Kenya | A | 2396.3 | 0.99 | 4.4 |
| South Sudan | A | 242.5 | 0.88 | 1.1 |
| China | A | 234.4 | 0.88 | 1.0 |
| Nigeria | A | 14.5 | 0.31 | 0.4 |
| Denmark | A | 7.6 | 0.19 | 0.2 |
| Italy | B.1 | 657.2 | 0.95 | 1.6 |
| Uganda | B.1 | 469.0 | 0.93 | 1.5 |
| Kenya | B.1 | 208.4 | 0.85 | 3.2 |
| Tanzania | B.1 | 195.1 | 0.84 | 1.2 |
| Morocco | B.1 | 156.5 | 0.81 | 1.1 |
| Democratic Republic of the Congo | B.1 | 49.7 | 0.58 | 1.1 |

**Supplementary Figure S1.**
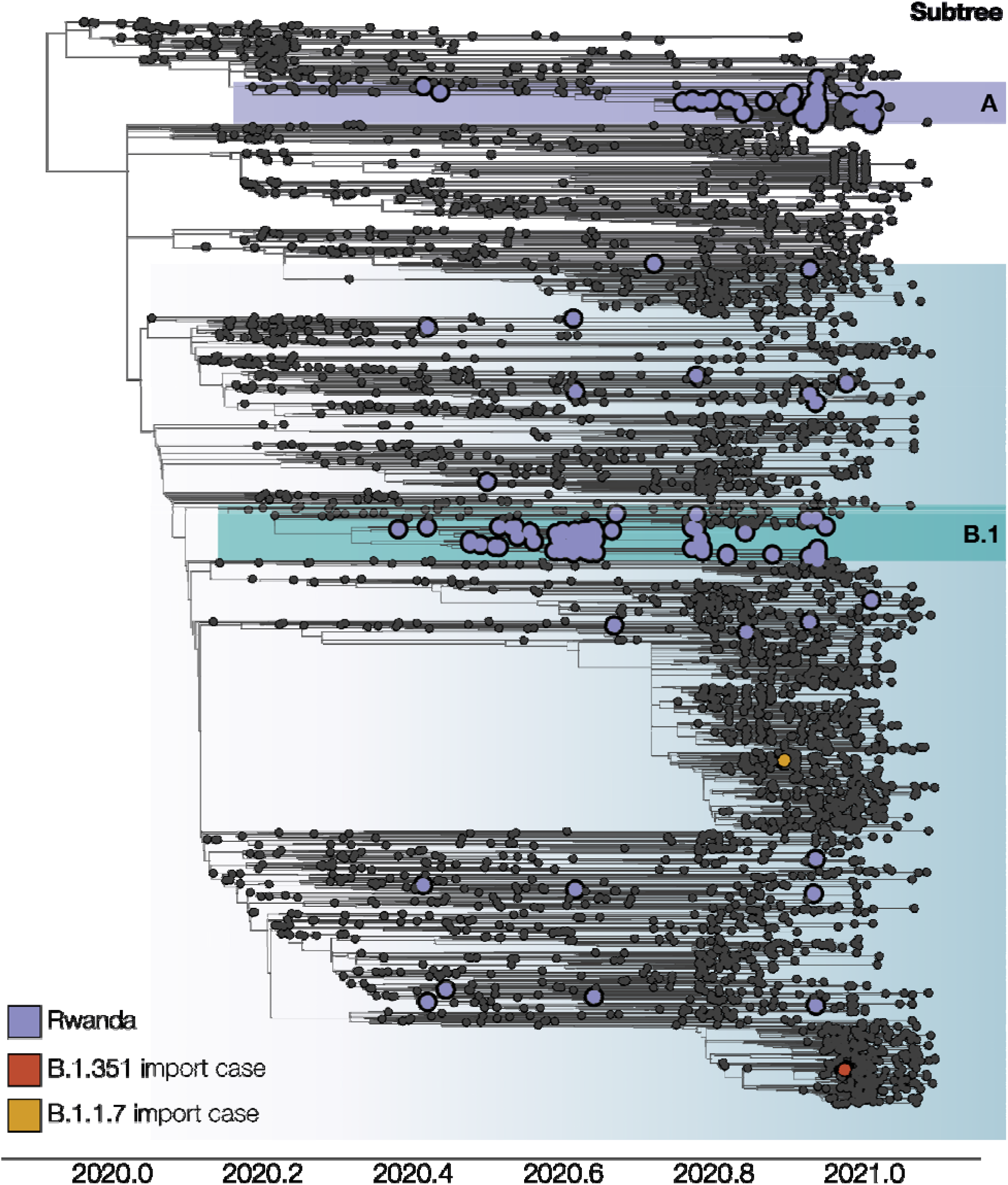
Time-calibrated phylogenetic tree of Rwandan SARS-CoV-2 genome sequences in the global context of the current SARS-CoV-2 pandemic. Two large clusters of Rwandan sequences can be identified, representing lineages A.23.1 and B.1.380, with other individual Rwandan sequences scattered throughout the phylogeny. Subtrees encompassing lineages A.23.1 and B.1.380 (referred to as subtree A and subtree B.1) were selected to perform Bayesian phylogeographic reconstruction that accommodates individual travel histories. Two variants of concern (VOC) sequences were detected during sequencing: one from lineage B.1.1.7, a returning traveler from Burundi, and one from lineage B.1.351, a returning traveler from the Democratic Republic of the Congo.

**Supplementary Figure S2.**
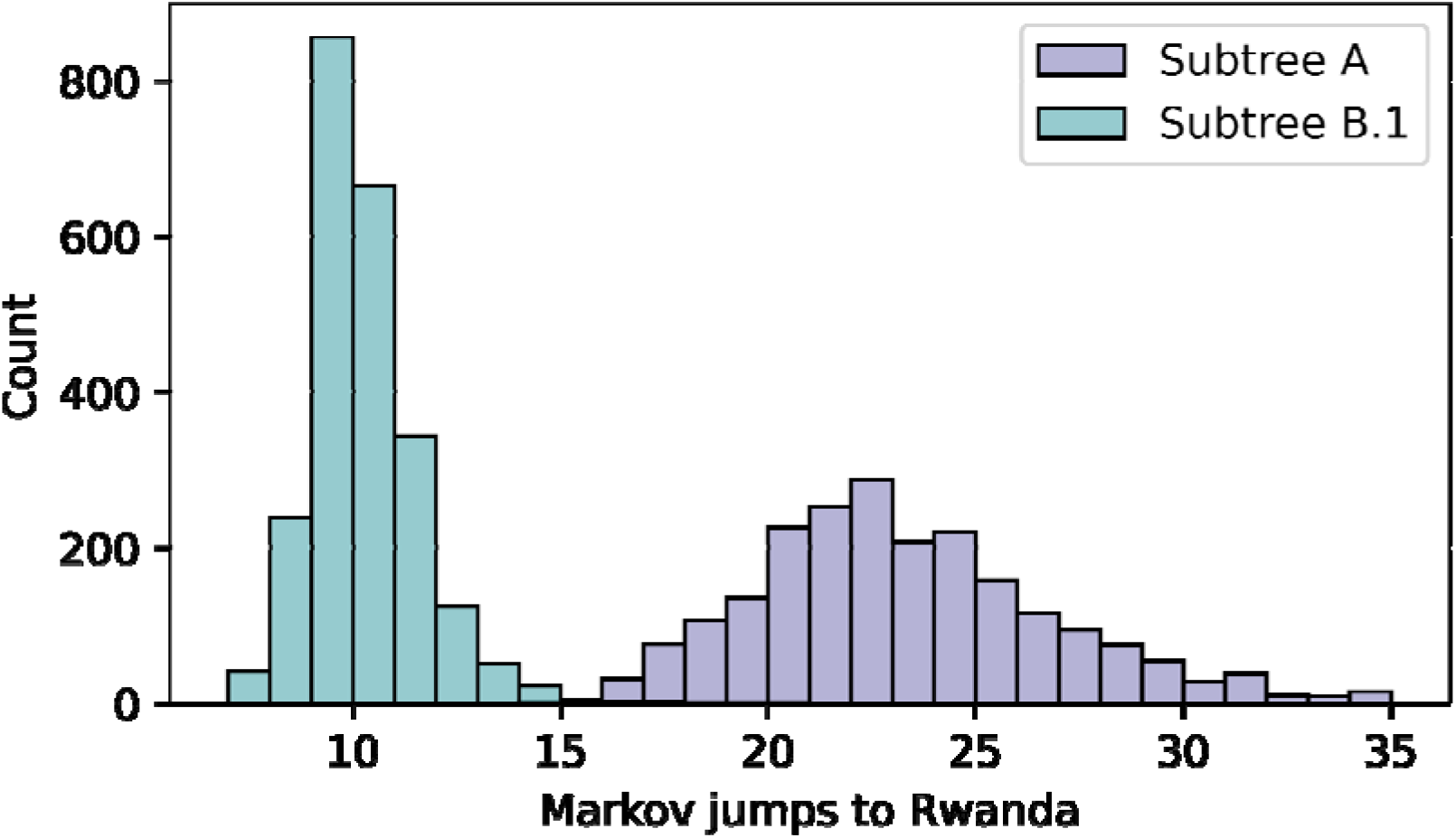
Posterior number of introductions into Rwanda for subtrees A and B.1. The total number of Markov jumps into Rwanda for each subtree was estimated via stochastic mapping on an asymmetric discrete state phylogeographic model. Despite the lower number of Rwandan sequences (subtree A: 49; subtree B: 134), subtree A reveals a higher number of introduction events (mean=22.8; 95%HPD=[16-29]) compared to subtree B.1 (mean=9.8; 95%HPD=[8-12]).

**Supplementary Figure S3.**
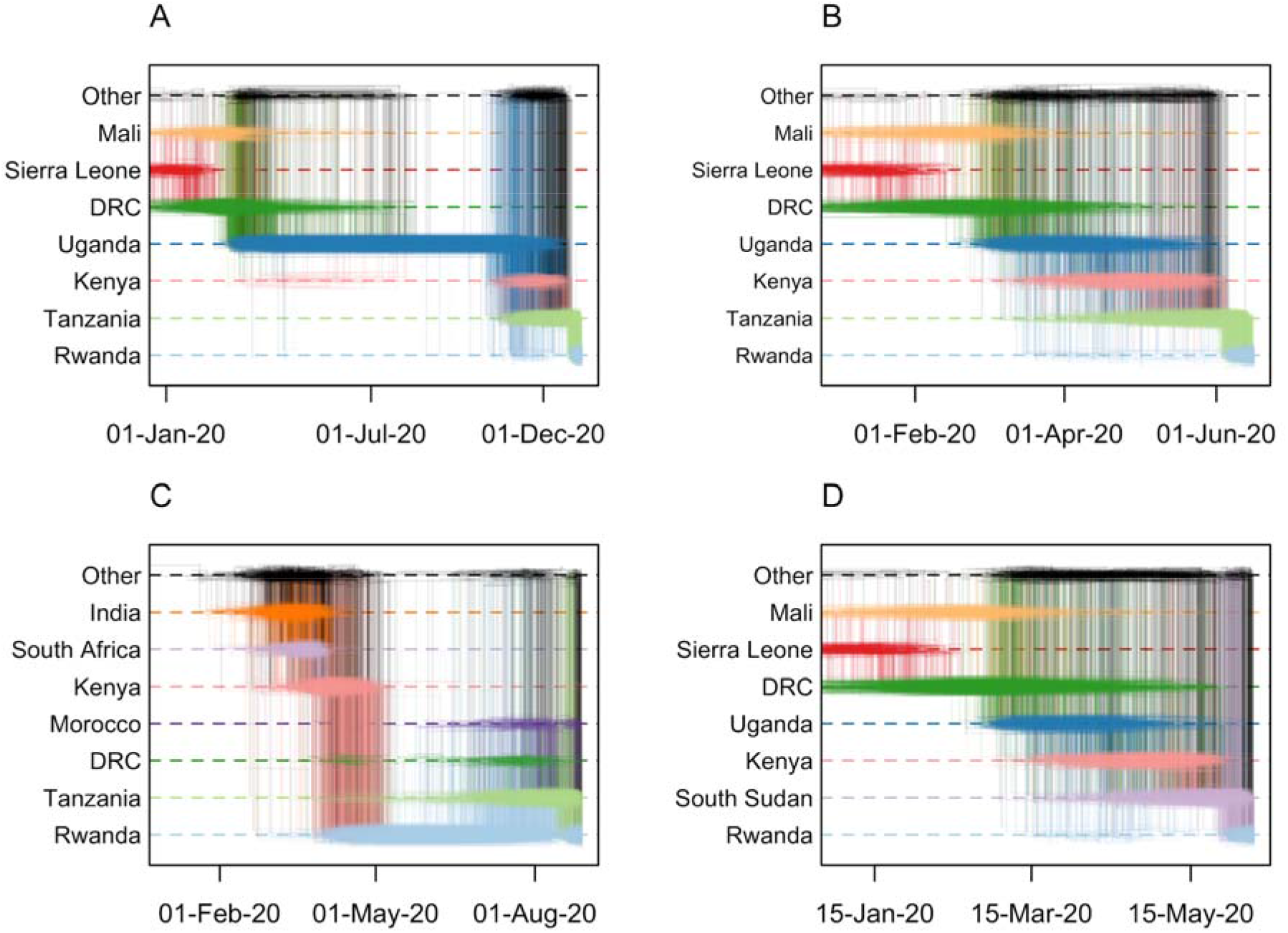
Markov jump trajectory plots for four selected Rwandan infected individuals with travel history (returning) from Tanzania (A, B, C) and South Sudan (D). Each individual trajectory corresponds to the Markov jumps in a single tree from the posterior distribution, with each plot showing the uncertainty across a subsample of 1,000 posterior trees. The horizontal dimension represents the time maintained at an ancestral location. Vertical lines represent a Markov jump between two locations. The seven most prominent locations across all ancestral paths in the posterior are displayed along the Y-axis, with “Other” representing the remaining locations. Trajectory plots A, B and D correspond to the isolates in subtree A, i.e. EPI_ISL_1064164, EPI_ISL_707772, and EPI_ISL_707712 respectively. Trajectory plot C corresponds to isolate EPI_ISL_960250 in subtree B.1. In all cases, considerable uncertainty in the ancestral reconstructions can be seen from the pattern of overlapping horizontal lines and the diffuse density of vertical lines, which indicate considerable support for different ancestral locations (i.e. uncertainty in the spatial reconstruction), and variance in the reconstructed timing of the introductions. For trajectory plots A, B and D, we observe similar patterns in the spatial paths reconstructed, where the isolates find ancestries in Mali / Sierra Leone / Democractic Republic of Congo, Uganda and Kenya prior to each corresponding travel location. In contrast, trajectory plot C shows support for an ancestry in Rwanda prior to the virus circulating in Tanzania and being reintroduced into Rwanda. This indicates a bidirectional exchange of viral lineages between the two countries, although the possibility of an intermediary country being involved cannot be discarded due to unevenness in sampling efforts between countries.

**Supplementary Figure S4.**
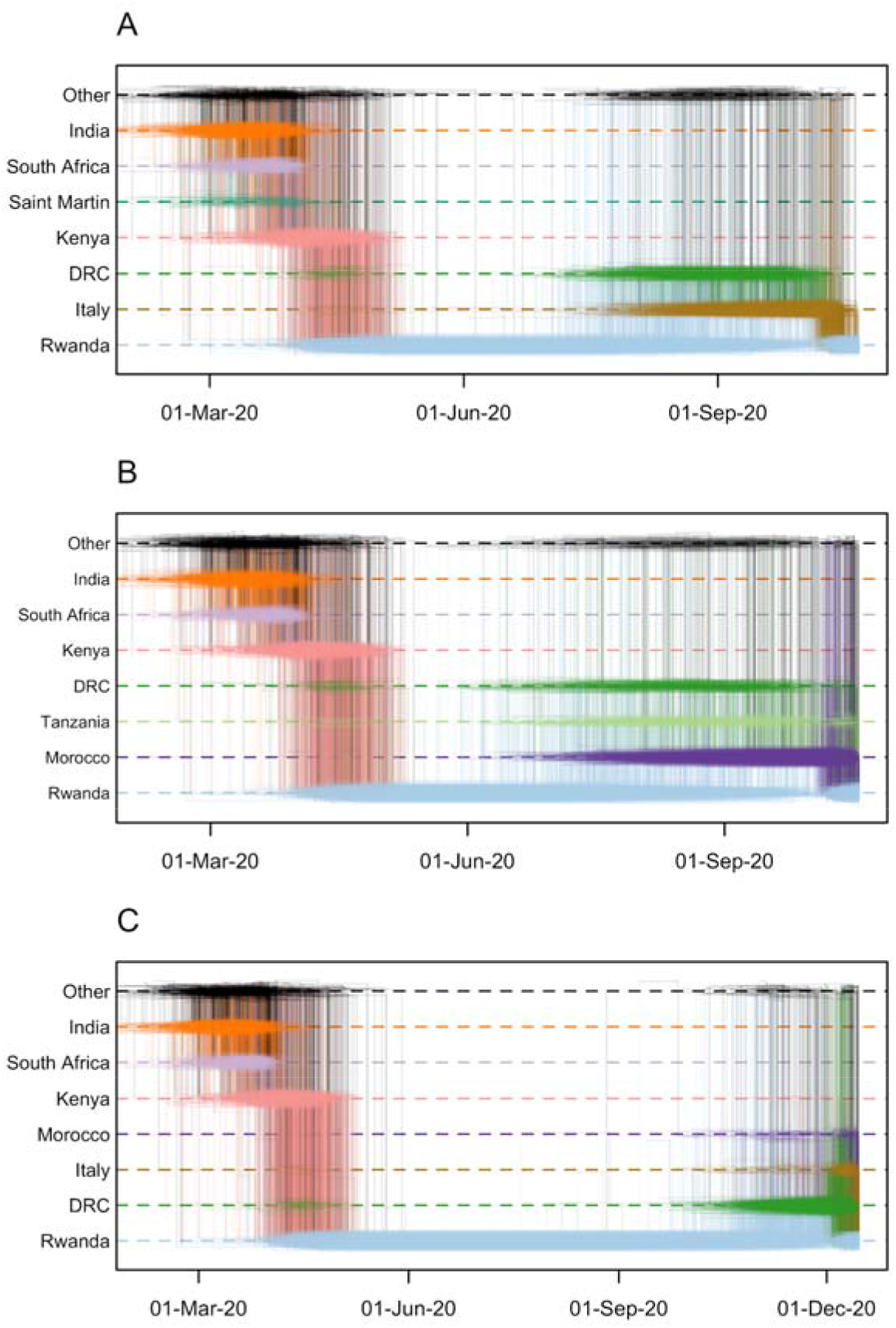
Markov jump trajectory plots for three selected Rwandan infected individuals with travel history (returning) from Italy (A), Morocco (B) and the Democratic Republic of the Congo (C). Similar to Figure S3 C, the ancestral histories inferred for these three isolates show support for a bidirectional flow of viral lineages between each corresponding travel location and Rwanda.

**Supplementary Figure S5.**
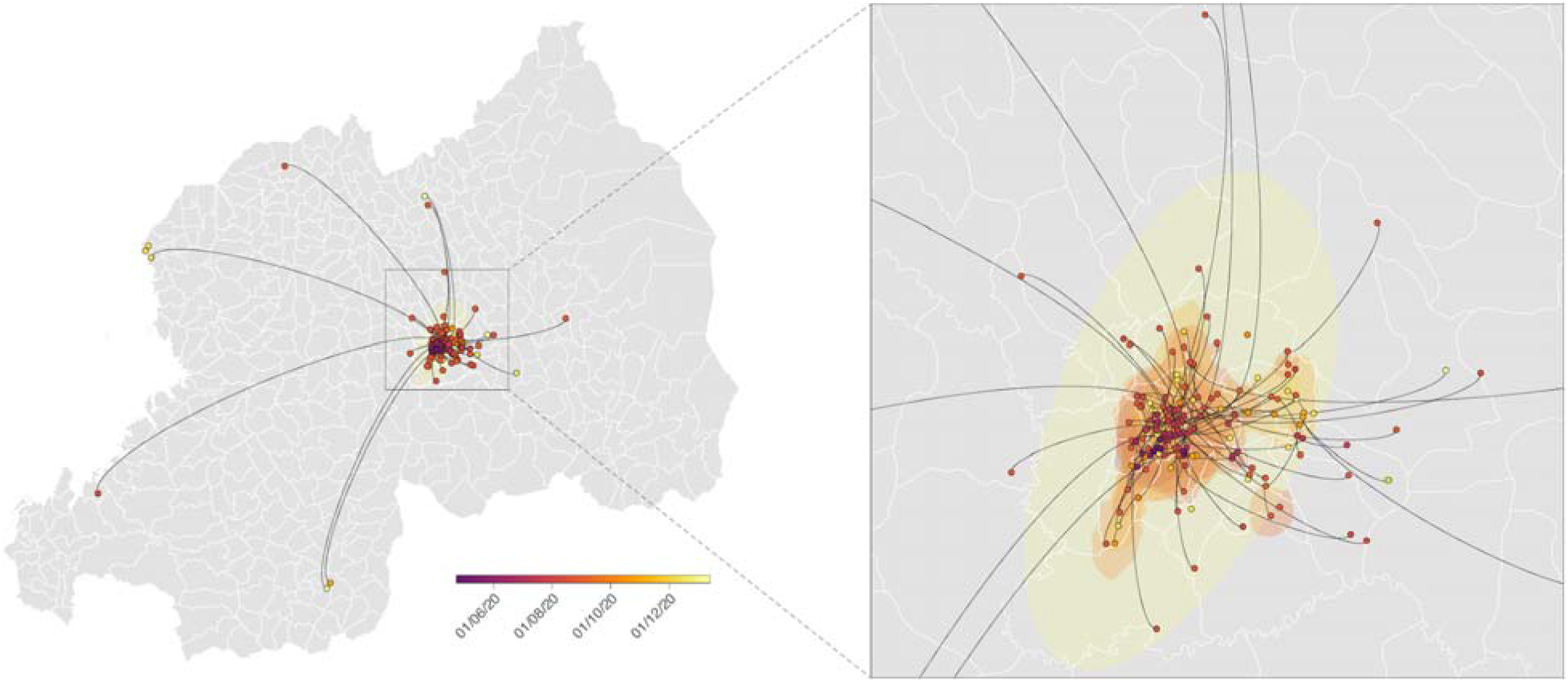
Spatially-explicit phylogeographic reconstruction of the dispersal history of SARS-CoV-2 lineages sampled in Rwanda. Spatially-explicit phylogeographic reconstruction was performed along the Rwandan clades identified within the two subtrees A and B.1. For each clade we mapped the maximum clade credibility (MCC) tree and overall, 80% highest posterior density (HPD) regions reflecting the uncertainty related to the phylogeographic inference. MCC trees and 80% HPD regions are based on 1,000 trees subsampled from each post burn-in posterior distribution. MCC tree nodes were colored according to their time of occurrence, and 80% HPD regions were computed for successive time layers and then superimposed using the same color scale reflecting time. Continuous phylogeographic reconstructions were only performed along clades linking at least three sequences sampled in Rwanda and for which the sector of origin was known. Besides the phylogenetic branches of MCC trees obtained by continuous phylogeographic inference, we also mapped sampled sequences belonging to clades linking less than three geo-referenced sequences. Sector borders are represented by white lines.

